# Intramedullary Lesions and Perilesional Tissue Bridges Predict Motor Recovery in Degenerative Cervical Myelopathy: A Multicenter Study

**DOI:** 10.1101/2025.08.05.25333069

**Authors:** Fauziyya Muhammad, Dario Pfyffer, Todd Parrish, Serena S. Hu, John Ratliff, Nathaniel D. Stetson, Yasin Dhaher, Andrew Smith, Kenneth A. Weber, Zachary A. Smith

## Abstract

**Background and Objectives:** Degenerative cervical myelopathy (DCM) is the leading cause of spinal cord-related disability worldwide. T2-weighted (T2w) intramedullary hyperintensities are common MRI findings in DCM and may reflect irreversible pathology, namely intramedullary lesions. The prognostic value of both quantitative lesion characteristics and the integrity of surrounding spared spinal cord tissue—tissue bridges—remains unclear. Using automated quantification of intramedullary lesions and tissue bridges, this study aimed to determine whether these structures can serve as imaging biomarkers of clinical severity and predict functional recovery in DCM.

**Methods:** This retrospective, multicenter study included 94 patients with DCM from four tertiary centers. All underwent baseline MRI and clinical assessment, with 6-month follow-up in 48.9% (n=46) of patients at six months. We used SCIsegV2, an automated tool integrated in the Spinal Cord Toolbox, to segment T2w hyperintense lesions and midsagittal tissue bridges. Lesion characteristics (volume, length, width, maximal axial damage ratio) and tissue bridge widths were extracted. Clinical assessments included mJOA, Neck Disability Index (NDI), hand dexterity, and balance. We evaluated associations between imaging metrics and clinical outcomes by using correlation and multivariable linear regression models adjusted for baseline clinical score, age, sex, and surgical status.

**Results:** Intramedullary lesions were present in 45.7% (n=43) of patients and were associated with greater clinical severity. Lesion volume, length, and maximal axial damage ratio (MADR) correlated with dexterity and balance impairments, but not with mJOA or NDI. Wider tissue bridges were associated with better dexterity at both time points. Multivariable models showed lesion volume and MADR independently predicted poorer balance at follow-up, while tissue bridge width was positively linked to with dexterity improvements. In surgical patients, lesion magnitude and tissue bridge integrity explained up to 71% of the variance in follow-up outcomes.

**Discussion:** Automated quantification of intramedullary lesion extent and spared tissue bridges provide robust biomarkers for structural damage and preserved function in DCM. These features better predict recovery, particularly in dexterity and balance, than conventional metrics. Integrating these metrics into clinical workflows could enhance surgical decision-making and support personalized prognosis. Future studies should incorporate 3D segmentation and multimodal imaging to refine predictions of long-term outcomes prediction.

## INTRODUCTION

T2-weighted (T2w) magnetic resonance imaging (MRI) plays a pivotal role in evaluating spinal cord pathology in degenerative cervical myelopathy (DCM)^1–3^. Hyperintense intramedullary lesions on T2w imaging may reflect irreversible pathological changes, such as ischemia, gliosis, demyelination, and necrosis^4,5^. While smaller or absent lesions are generally associated with better functional outcomes, the prognostic value of lesion extent in chronic compressive myelopathy remains underexplored, especially in contrast to similar findings in acute traumatic spinal cord injury (SCI). The structural integrity of spared spinal cord tissue adjacent to intramedullary lesions, typically referred to as ‘tissue bridges’, has emerged as a critical predictor of neurological and functional recovery in SCI^6–8^. These bridges cover preserved axonal pathways and are thought to facilitate and mediate residual motor and sensory conduction^8,9^.

Recent advancements in quantitative neuroimaging and open-source analysis tools have significantly improved the reliability and granularity of spinal cord lesion assessments. Notably, the automated lesion segmentation tool SCIsegV2, developed as part of the Spinal Cord Toolbox (SCT), allows for accurate 3D quantification of T2w hyperintense lesions and tissue bridges^10,11^. Its validation in multi-site SCI studies positions it as a promising solution to overcome the limitations of manual lesion annotation, including observer variability, image-plane dependency, and limited reproducibility.

In this study, we apply the automated quantification of lesions and tissue bridges via SCIsegV2 to a large, multicenter cohort of patients with DCM to systematically evaluate the relationship between intramedullary lesion and tissue bridge parameters and post-surgical outcomes. Our goal is to establish whether automated quantification of T2w hyperintense lesions and spared tissue bridges can serve as robust, imaging-derived biomarkers of disease severity and predictors of functional recovery in chronic cervical spinal cord compression. We hypothesize that reduced intramedullary lesion burden and preserved tissue bridge integrity, quantified via automated segmentation, will be significantly associated with improved functional outcomes in DCM patients. This work advances the application of structural MRI from descriptive pathology toward precision prognostication in DCM.

## METHODS

### Standard Protocol Approvals, Registrations, and Patient Consents

This retrospective, longitudinal, multi-site study included patients diagnosed with DCM across four tertiary academic centers: the University of Oklahoma Health Sciences Center (OUHSC), Northwestern University, University of Texas Southwestern (UTSW), and Stanford University. Institutional Review Board (IRB) approval was obtained from each participating institution, including OUHSC (IRB #15513). All patients provided written informed consent.

### Participants and Experimental Design

A total of 94 patients with DCM were enrolled from the four academic institutions: OUHSC (n=52), Northwestern University (n=21), UTSW (n=10), and Stanford University (n=11). Patient recruitment and MRI/clinical examinations were conducted between April 2018 and May 2025, with site-specific timeframes as follows: OUHSC (5/21/2021–5/25/2025), Northwestern (4/12/2018–1/17/2020), UTSW (3/30/2022–8/17/2024), and Stanford (………..). All patients underwent baseline MRI and clinical assessments. Follow-up assessments were conducted six months post-surgery or, for non-surgical patients, six months after baseline evaluation. Inclusion criteria required a diagnosis of DCM and visibility of cervical spinal cord pathology on T2w imaging. Patients with thoracic-only lesions were excluded and those without intramedullary lesion were not included in the lesion analyses.

### Clinical Assessment

Clinical measures were collected as follows: mJOA scores in 94 patients, Neck Disability Index (NDI) in 84 patients (UTSW patients excluded), dexterity scores (NIH Toolbox Nine-Hole Peg Test) in 73 patients, and balance scores (postural sway analysis) in 69 patients^12,13^. Follow-up clinical scores were obtained at six months for 46 patients, including 32 who underwent surgical decompression and 14 who were managed non-surgically.

### MRI Acquisition

T2-weighted MRIs of the cervical spine were acquired at 3T on a Siemens scanner equipped with 16 channel spine coil (UTSW, Stanford, and Northwestern) or General Electric (GE) MR750w scanner equipped with 16 channel head-neck-spine coil (OUHSC). Two acquisition protocols were used, an isotropic resolution (0.8 × 0.8 × 0.8mm^3^) and with field of view (FOV) of 256 × 256, see generic spine protocol) for MRI at OUHSC, UTSW, and Stanford. And anisotropic resolution, 1 × 1 × 2mm^3^, FOV of 180 × 180 for MRI acquired at Northwestern. **Supplementary Table 1** provides detailed imaging parameters from each acquisition site. For additional protocol information, see the generic protocol and Paliwal *et al* for-acquisition details^14,15^.

**TABLE 1:** DEMOGRAPHIC INFORMATION OF PATIENTS WITH DCM.

| TABLE 1: DEMOGRAPHIC INFORMATION OF PATIENTS WITH DCM |  |  |  |  |  |
| --- | --- | --- | --- | --- | --- |
| SITES | OUHSC | NorthWestern | UTSW | Stanford | Total |
| Participants (n) | 52 | 21 | 10 | 11 | 94 |
| F/M | 29/23 | 9/12 | 5/5 | 6/5 | 49/45 |
| Mean Age(SD) | 54.40 (9.31) | 58.05 (13.56) | 60.8 (13.79) | 54 (11.79) | 55.85 (11.21) |
| Ave. mJOA (SD) | 13.88 (2.57) | 14.29 (1.79) | 14.6 (1.72) | 14.36 (2.16) | 14.11 (2.27) |
| **Disease Severity** |  |  |  |  |  |
| Mild (15-17) | 24 | 9 | 8 | 7 | 48 |
| Moderate (12-14) | 17 | 12 | 1 | 3 | 33 |
| Severe (<11) | 11 | 0 | 1 | 1 | 13 |
| **Follow up scores (n)** | 33 | 13 | 0 | 0 | 46 |
| Surgery | 19 | 13 | 0 | 0 | 32 |
| Non-surgery | 14 | 0 | 0 | 0 | 14 |
| **Intramedullary Lesion Present** |  |  |  |  |  |
| Yes | 21 | 8 | 7 | 7 | 43 |
| No | 31 | 13 | 3 | 4 | 51 |
| **Clinical Assessment Scores (n)** |  |  |  |  |  |
| mJOA | 52 | 21 | 10 | 11 | 94 |
| NDI | 52 | 21 | 0 | 11 | 84 |
| Balance | 49 | 0 | 9 | 11 | 69 |
| Dexterity | 52 | 0 | 10 | 11 | 73 |

For image preprocessing, all DICOM images were converted to NifTi, reoriented to RPI, and rigidly registered for quality check (QC) and spinal cord segmentation. All T2-weighted images were resampled to an in-plane resolution of 0.6 mm x 0.6 mm in the sagittal plan to optimize spinal cord segmentation using the SCT’s deep-learning algorithms (SCT v7). Spinal cord segmentation masks were then non-linearly registered to PAM50 spinal cord template (1mm x 1 mm x 1 mm) for anatomical normalization.

### Image Analysis

T2w hyperintense intramedullary lesions were confirmed in 43 patients; the remaining 51 patients had no visible cervical cord lesion at baseline. Automated segmentation and characterization of intramedullary lesions and midsagittal tissue bridges were performed using the SCIsegV2 tool, integrated into SCT (v7). Lesion metrics included rostrocaudal lesion length, dorsoventral lesion width, maximal axial damage ratio, and the width of preserved tissue bridges measured as the sum of ventrally and dorsally spared regions flanking the lesion. All measures were obtained from the mid-sagittal slice to ensure consistency and due to their prognostic value reported previously in traumatic SCI^7,8^. Visual depictions are presented in **Figure 1A–1E**.

**Figure 1.**
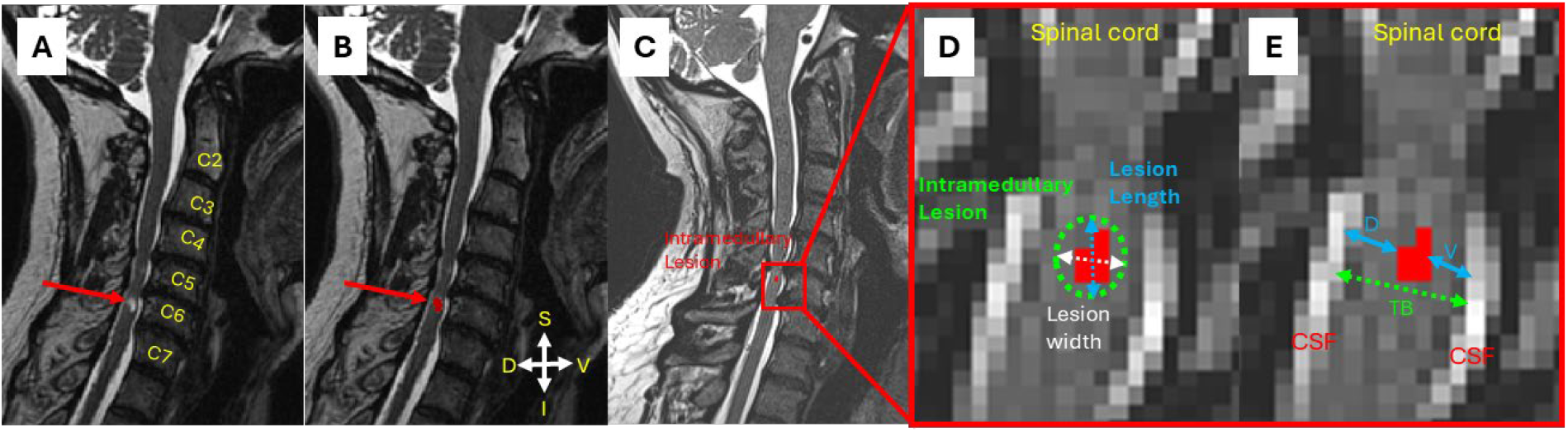
Schematic of intramedullary lesion segmentation and characterization in a T2-weighted cervical spine MRI. Panels A–E illustrate the key structural features used for quantitative lesion characterization, including lesion dimensions and preserved spinal cord tissue around the lesion (bridges). A. Representative sagittal T2-weighted MRI from a patient with degenerative cervical myelopathy (DCM), illustrating a hyperintense intramedullary lesion at the C6 vertebral level. B. Automated lesion segmentation using the Spinal Cord Toolbox (SCT), applying the C6-specific algorithm (version 2) for accurate identification and localization. C. Magnified view of the lesion for detailed assessment, demonstrating spatial orientation (S: superior, I: inferior, D: dorsal, V: ventral) and lesion boundary. D. Axial view highlighting key lesion metrics: lesion width (green dotted line) and lesion length (blue line). E. Tissue bridge analysis in the axial plane showing spared dorsal (D) and ventral (V) tissue bridges (TB, green line) surrounding the lesion (red), with surrounding cerebrospinal fluid (CSF) visible.

### Statistical Analysis

All statistical analyses were performed using GraphPad Prism version 10.3.1 (GraphPad Software, La Jolla, CA) and Python 3.9 libraries (Pandas, Matplotlib, Seaborn, Numpy, and SciPy Stats; Python Software Foundation, Python.org). The distribution of continuous variables was assessed using the D’Agostino-Pearson omnibus normality test using GraphPad Prism. Depending on normality, Pearson or Spearman correlation coefficients were used to evaluate associations between lesion features (volume, length, width, axial damage ratio, total bridge width) and clinical measures (mJOA, NDI, dexterity, balance). Kruskal-Wallis tests followed by Dunn’s multiple comparison tests were applied for group-wise comparisons of lesion and bridge characteristics across DCM severity categories and bridge size subgroups. Categorical data, such as surgical rates by lesion status, were compared using chi-square tests.

To evaluate the association between lesion features on post-surgical functional recovery, multivariable linear regression models were constructed. These models adjusted for baseline clinical scores, age, sex, and surgical status. Results are reported as beta coefficients (β) with corresponding model R^2^ values. P-values reported for secondary and exploratory analyses are nominal and were not corrected for multiple comparisons.

### Data Availability Statement

All analysis code and sample data used for lesion and tissue bridge quantification are available via a public GitHub repository (https://github.com/Mfauziyya/Tissue-Bridges). The full dataset supporting the findings of this study is available from the corresponding author upon reasonable request, pending IRB and institutional data sharing policies.

## RESULTS

### Demographic and Clinical Characteristics

Ninety-four patients with DCM were enrolled across four tertiary care centers. The DCM cohort had a mean age of 55.9 ± 11.2 years (range: 25–80), with 52.1% females (n = 49). The mean mJOA score for all participants was 14.1 ± 2.3, and we stratified them based on disease severity into mild (mJOA 15–17; n = 48), moderate (mJOA 12–14; n = 33), and severe (mJOA < 12; n = 13) groups. There was no significant difference in the distribution of DCM severity across recruitment sites (χ^2^ = 2.14, p = 0.541). **Table 1** summarizes demographic data for the patients included in this study.

### Association of Intramedullary Lesion Prevalence and Extent with DCM Severity

A total of 43 of 94 patients (45.7%) demonstrated T2w hyperintense intramedullary lesions at or near the level of maximal compression (MCL) (**Figure 1A**). Lesion prevalence increased with clinical severity with 43.8% in mild, 45.5% in moderate, and 53.8% in severe DCM cases, although these differences were not statistically significant (Fisher’s exact test, χ^2^ = 0.82, p = 0.660). Most patients had a single lesion (n = 27), with a mean of 1.58 ± 0.7 lesions per patient. Patients with severe DCM had a slightly higher number of lesions per patient (2.0 ± 0.8) compared those with moderate (1.73 ± 0.6) or mild (1.33 ± 0.5) DCM; however, these differences were not statistically significant (**Table 2**).

**TABLE 2:** SUMMARY STATISTICS OF LESION (n=43) BY mJOA CATEGORY.

| **TABLE 2: SUMMARY STATISTICS OF LESION (n=43) BY mJOA CATEGORY** |  |  |  |
| --- | --- | --- | --- |
| mJOA Category | # of patients | % of severity group | Ave. # of lesions per subject |
| Mild (15-17) | 21 | 43.75 | 1.33 |
| Moderate (12-14) | 15 | 45.45 | 1.73 |
| Severe (<11) | 7 | 53.84 | 2 |
| Total | 43 | 45.74 | 1.58 |

We examined whether lesion characteristics were associated with DCM severity. Lesion volume significantly differed across severity groups (Kruskal–Wallis test, p = 0.015), with the highest mean volume in severe DCM (234.4 ± 212.1 mm^3^), compared mild (79.0 ± 84.5 mm^3^, p = 0.010) or moderate (61.6 ± 85.6 mm^3^, p = 0.007) DCM (**Figure 2A–D**). Participants suffering from severe DCM (12.6 ± 7.2 mm) had significantly longer lesions than those with mild (6.4 ± 3.1 mm, p = 0.023) but not moderate disease (6.9 ± 4.7 mm, p = 0.078). No significant differences were observed in lesion width between severity groups (Kruskal–Wallis test, *p* = 0.194). The maximal axial damage ratio defined as the cross-sectional area of the lesion relative to total cord area was significantly greater in severe DCM (0.5 ± 0.2) than in moderate (0.2 ± 0.2, p = 0.004) or mild (0.3 ± 0.0, p = 0.020) cases (Kruskal–Wallis test, *p* = 0.021). These findings suggest that intramedullary lesions are associated with a more severe clinical presentation.

**Figure 2.**
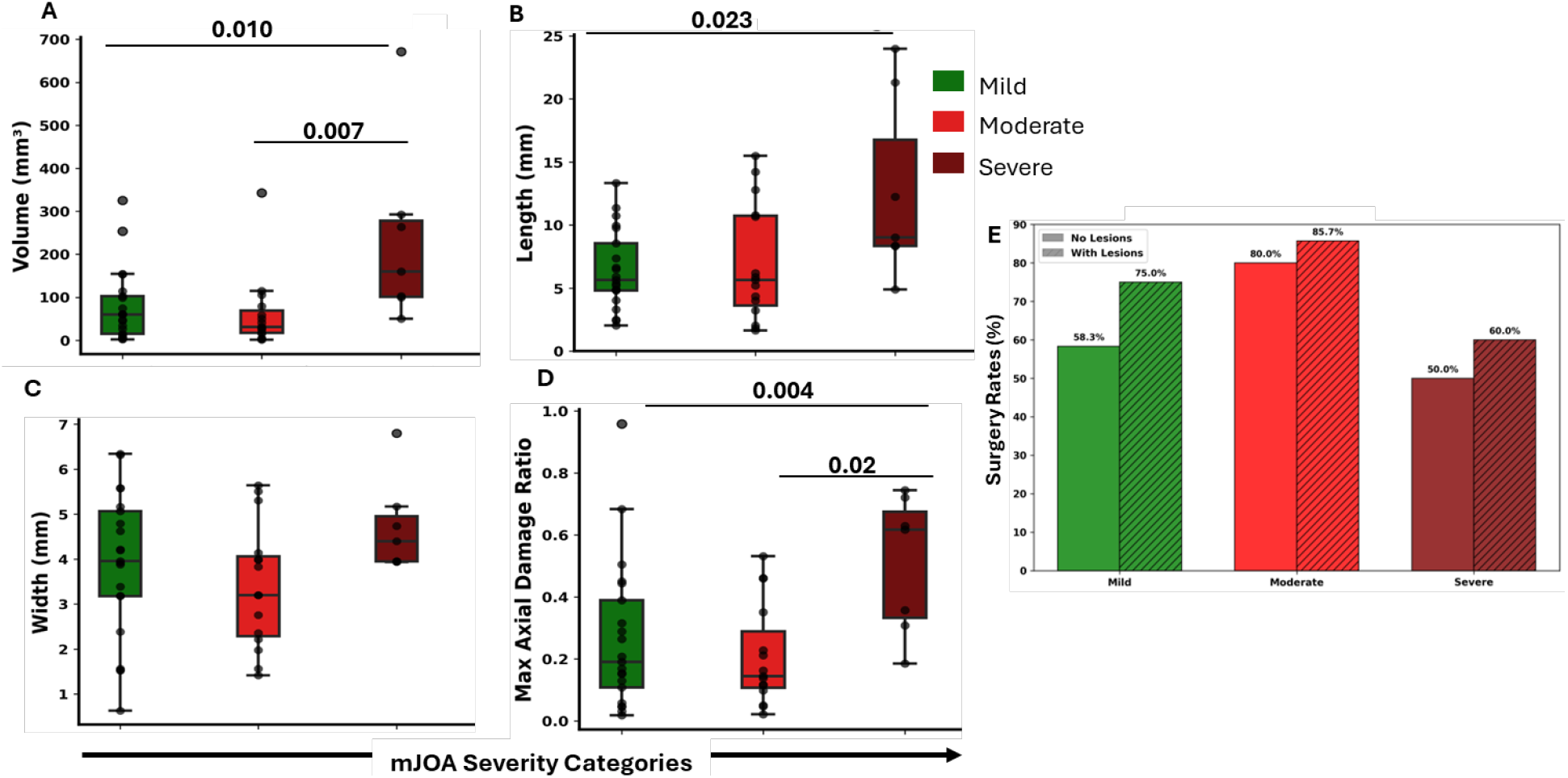
Association between lesion characteristics and disease severity in degenerative cervical myelopathy (DCM). A–D. Box plots showing lesion metrics stratified by mJOA severity categories (mild, moderate, severe). A. Lesion volume (mm3), B. Lesion length (mm), C. Lesion width, and D. Maximal axial damage ratio were significantly higher in patients with severe myelopathy, as determined by Mann-Whitney test with Bonferroni correction (p-values indicated). Lesion volume, length, and maximal axial damage ratio significantly differed across severity groups. E. Bar graph comparing surgery rates between patients with and without intramedullary lesions across severity strata. The presence of a lesion was associated with higher surgical rates in all severity groups.

### Influence of Intramedullary Lesions on Surgical Decision-Making

To assess whether the presence of lesions influenced surgical decision-making, we analyzed data from 46 patients with DCM who had a known surgical status. Among patients without a lesion (n = 26), the surgical rate was 65.4%, whereas the rate among those with a lesion was 75.0%; however, this difference was not statistically significant (Fisher’s exact test, *p* = 0.535). When stratified by severity, mild cases of DCM in which lesions were present appeared to influence surgical decisions most. In mild cases, surgery rates rose from 58.3% (no lesion) to 75.0% (with lesion); in moderate cases, 80.0% (no lesion) vs. 85.7% (with lesion); and in severe cases, 50.0% (no lesion) vs. 60.0% (with lesion); these differences are not statistically significant (**Figure 2E**. These findings suggest that lesion detection may prompt earlier surgical intervention, especially in patients with mild symptoms. It is possible that those with more severe cases of DCM also have a greater number of contraindications or comorbidities than those with less severe cases. If this is the case, then DCM severity, rather than the presence of lesions per se, could be a key determining factor in the surgical decision-making process and account for the comparatively lower surgical rate observed in this group. Overall, lesion presence may serve as a surrogate for disease burden and inform surgical decision-making in milder cases.

### Association of Lesion Characteristics with Functional Deficits in DCM

We next evaluated the relationship between lesion characteristics (volume, length, width, maximal axial damage ratio) and clinical impairment, including mJOA, NDI, hand dexterity, and balance. Balance performance was significantly negatively correlated with lesion volume (r = −0.45, p = 0.008), lesion length (r = −0.47, p = 0.006), and maximal axial damage ratio (r = −0.45, p = 0.009), and lesion width trended toward a negative association with balance performance (r = −0.30, p = 0.085). Dexterity also showed a significant negative correlation with the maximal axial damage ratio (r = −0.36, p = 0.032) and trended toward negative associations with lesion volume (r = −0.33, p = 0.053) and lesion length (r = −0.31, p = 0.075). No significant associations were found between lesion measures and mJOA or NDI after correction for multiple comparisons (**Figure 3**. These results suggest that larger lesions and greater axial involvement are linked to greater functional impairment.

**Figure 3.**
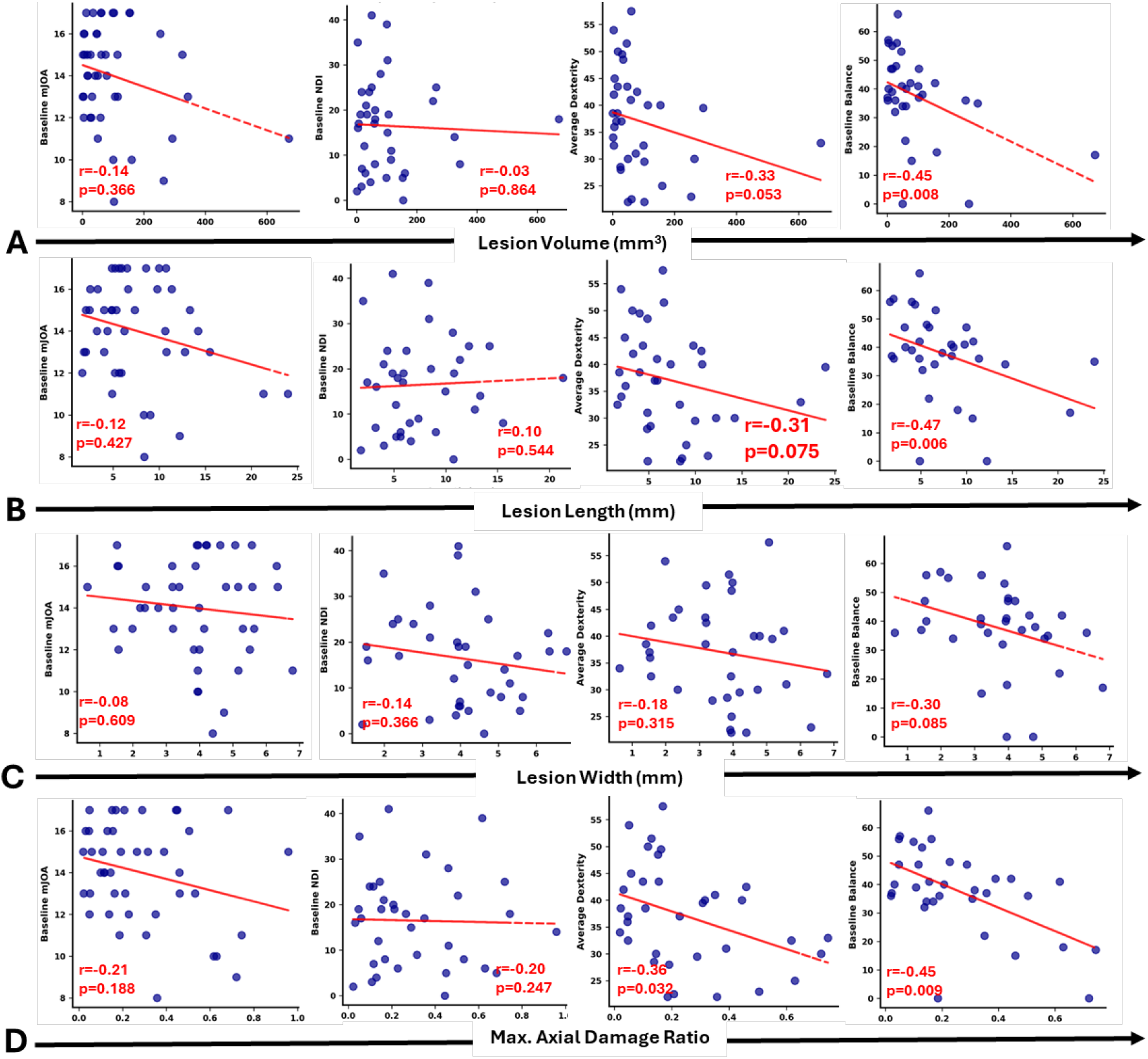
Association between lesion characteristics and baseline clinical scores in DCM. Each column of scatter plots shows the relationship between lesion measures and four baseline clinical scores: modified Japanese Orthopedic Association (mJOA), Neck Disability Index (NDI), dexterity, and balance. Lesion characteristics analyzed include: (A) Lesion volume, (B) Lesion length, (C) Lesion width, and (D) Maximal axial damage ratio. Lesion volume and length showed a negative correlation with dexterity and balance, with lesion volume significantly associated with baseline balance. Lesion width trended toward a negative association with balance. Maximal axial damage ratio significantly correlates with both dexterity and balance. No significant associations were observed between lesion characteristics and mJOA or NDI. Red lines denote linear regression fit.

### Preserved Tissue Bridge Widths across DCM Severity Categories

We measured the width of preserved spinal cord tissue (i.e., tissue bridges; **Figure 1E**) in patients with (n = 43) and without lesions (n = 51). For lesion-positive cases, dorsal, ventral, and total bridge widths were measured at the lesion site. For lesion-negative cases, the anterior– posterior (AP) diameter at the MCL was used as a surrogate. Across the mJOA severity categories, there were no significant differences in dorsal, ventral, or total tissue bridge width among patients with lesions (all *p* > 0.05) (**Figure 4A**). Tissue bridge lengths (i.e., dorsal, ventral, and total length) across DCM severity categories were as follows: Dorsal tissue bridge length in mild DCM was 0.5 ± 0.7 mm; moderate, 0.5 ± 0.7 mm; and severe, 0.7 ± 0.7 mm (Kruskal–Wallis test p = 0.569); ventral tissue bridge length in mild DCM was 0.6 ± 0.7 mm; moderate, 0.5 ± 0.6 mm; and severe, 0.6 ± 0.7 mm (Kruskal–Wallis test p = 0.874); total tissue bridge length in mild DCM was 1.1 ± 1.3 mm; moderate, 1.0 ± 1.2 mm; and severe, 1.3 ± 0.9 mm (Kruskal–Wallis test p = 0.542).

**Figure 4.**
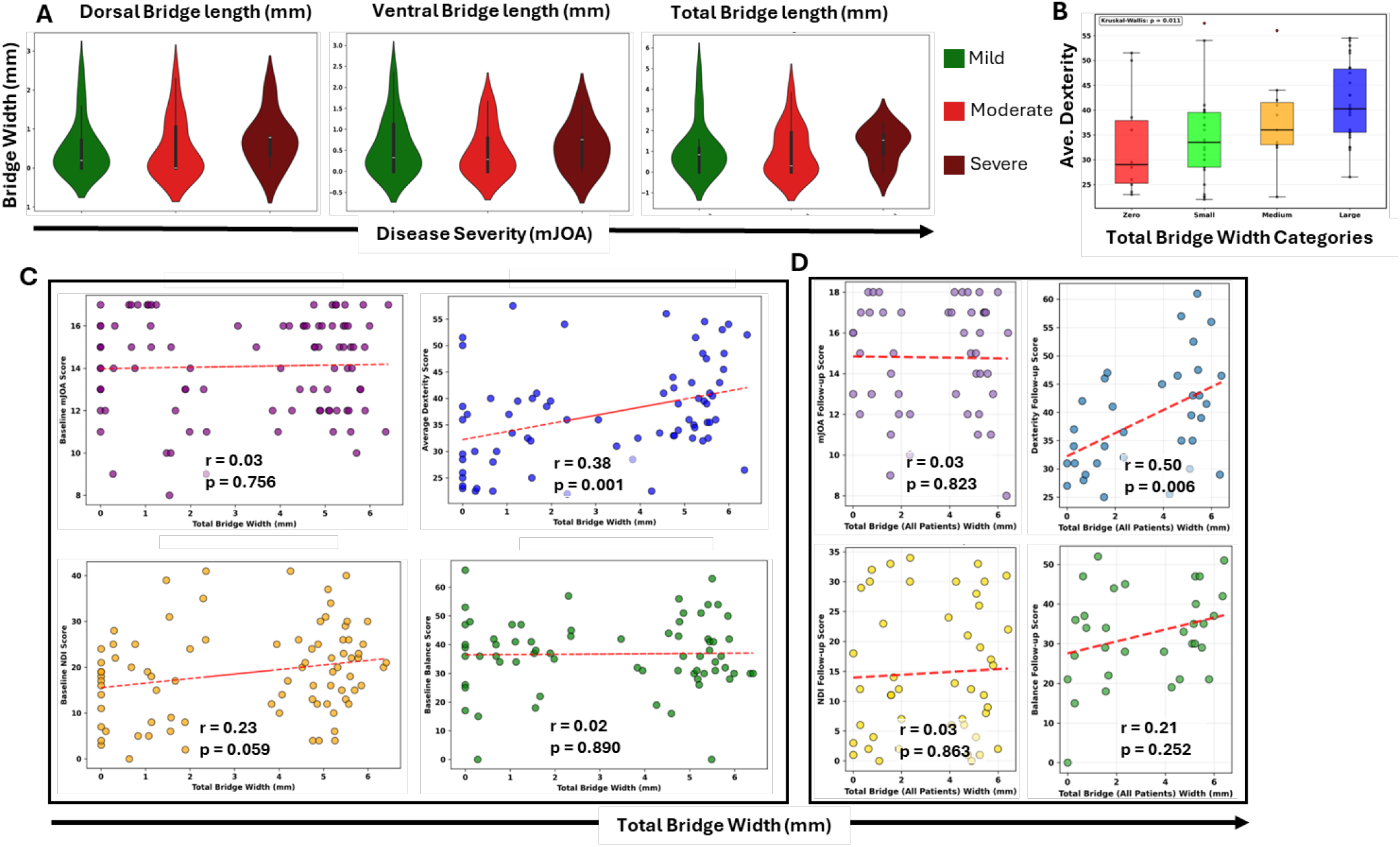
Association of preserved spinal cord tissue bridges with clinical outcomes in DCM. A. Violin plots illustrate the distribution of dorsal, ventral, and total tissue bridge widths (mm) across DCM severity categories (mild, moderate, severe). No significant differences were observed in bridge widths by severity. (B) Average dexterity scores demonstrated a significant group difference (p = 0.011). The Large bridge group had the highest mean ± s.d. dexterity score (41.58 ± 7.84). A progressive increase was observed across categories: Zero (33.15) → Small (34.18) → Medium (37.50) → Large (41.42). Pairwise comparisons revealed significant differences between Zero vs. Large (p = 0.020) and Small vs. Large (p = 0.004). Group sample sizes: Zero (n = 13, 13.8%), Small (n = 27, 28.7%), Medium (n = 26, 27.7%), Large (n = 27, 28.7%). Zero - Bright Red, Small (<2.41 mm) - Bright Green, Medium (2.41mm to 5.18 mm) - Orange, Large (≥ 5.18 mm) – Blue. C. Scatter plots show the relationship between total bridge width and baseline clinical scores. Total bridge width was positively associated with baseline dexterity but not significantly correlated with mJOA, NDI, or balance. D. Scatter plots show follow-up clinical outcomes in relation to tissue bridge width. Total bridge width remained significantly associated with follow-up dexterity, whereas associations with mJOA, NDI, and balance were not significant. Red lines indicate linear regression fits.

We further investigated the clinical impact of bridge size by categorizing patients into four groups: ‘Zero’ (0 mm, n = 13), ‘Small’ (<2.41 mm, n = 27), ‘Medium’ (2.41mm to 5.18 mm, n = 26), and ‘Large’ (> 5.18 mm, n = 27) total tissue bridge width. Mean dexterity scores significantly differed across these groups (Kruskal-Wallis p = 0.011), with the large bridge group demonstrating the highest mean dexterity performance (41.4 ± 7.8 T score). Pairwise post-hoc comparisons revealed significant differences between Zero vs. Large (p = 0.020) and Small vs. Large (p = 0.004), indicating that greater tissue bridge width is associated with improved hand function (**Figure 4B**). No significant group differences were found in baseline mJOA (p = 0.997), NDI scores (p = 0.153), or balance (p = 0.834) (**Supplementary Figure 1**).

### Clinical Relevance of Preserved Tissue Bridges

We further assessed whether preserved tissue bridges at the lesion site correlated with clinical outcomes (**Figure 4C-D**). In lesion-positive patients, dorsal, ventral, and total tissue bridge widths were not significantly different across severity groups. Total tissue bridge width showed a significant positive association with dexterity (*r* = 0.38, *p* = 0.001), a trend with NDI (*r* = 0.23, *p* = 0.059) scores, and no association with mJOA (*r* = 0.03, *p* = 0.756) or balance (*r* = 0.02, *p* = 0.890) scores (**Figure 4C**). Notably, some patients had a complete absence of dorsal or ventral bridges or both suggesting complete loss of preserved cord structure at the lesion site. Among the 43 lesion-positive patients, 20 lacked dorsal bridges, 15 lacked ventral bridges, and 13 lacked both (i.e., total tissue bridge width = 0 mm). Although spatial patterns of tissue bridge loss did not significantly affect clinical scores, trends indicated slightly higher mean balance scores in patients with preserved dorsal tissue bridges (**Supplementary Figure 2**).

### Longitudinal Clinical Relevance of Preserved Tissue Bridges

To assess the prognostic value of tissue bridges, we examined their association with follow-up clinical outcomes in patients with DCM (**Figure 4D**). In patients with known follow-up scores, total tissue bridge width was significantly positively associated with follow-up dexterity (*r* = 0.50, *p* = 0.006), suggesting that greater preservation of spinal cord architecture may contribute to improvements in hand function over time. No statistically significant associations were found between total tissue bridge width and follow-up mJOA (*r* = 0.03, *p* = 0.823) or NDI scores (*r* = 0.03, *p* = 0.863), but positive trends were observed for balance scores (*r* = 0.21, *p* = 0.252), although this association was not significant. These findings suggest that total bridge width could reflect the preservation (or lack thereof) of sensorimotor pathways relevant to upper-limb function recovery. If this is the case, then total bridge width could serve as an important marker for individuals with DCM.

### Prognostic Value of Lesion Characteristics and Tissue Bridges on Follow-up Outcomes

To further understand the prognostic relevance of imaging biomarkers, we performed a multiple linear regression analysis to evaluate the relative contribution of lesion volume, maximal axial damage ratio, and total tissue bridge width to follow-up clinical outcomes **(Table 3**). In the full cohort, the model explained 43% of the variance in follow-up balance scores, with lesion volume (β = −5.41) and maximal axial damage ratio (β = −5.57) emerging as the strongest negative predictors. For dexterity, the model accounted for 18% of variance, with lesion volume (β = −2.25) and total tissue bridge width (β = 2.11) contributing modestly.

**TABLE 3:**
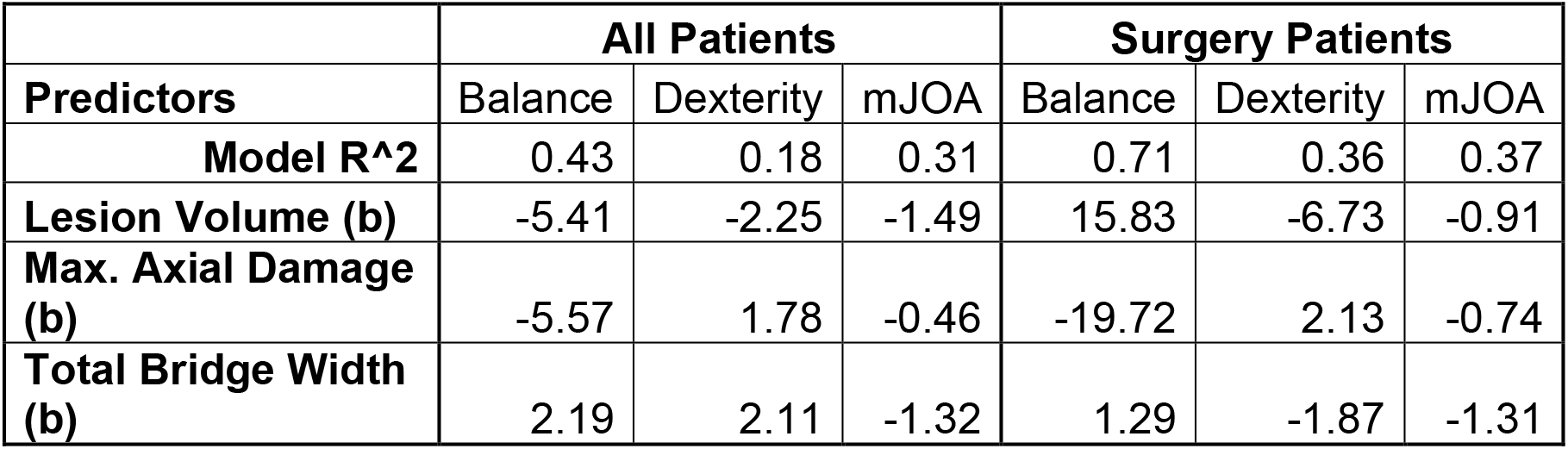
MULTIPLE REGRESSION ANALYSIS.

In patients who underwent surgery, the model was even more predictive for follow-up balance scores (R^2^ = 0.71), with maximal axial damage ratio showing a large negative association (β = −19.72), whereas lesion volume (β = 15.83) showed an unexpected positive trend. For post-surgical dexterity (R^2^ = 0.36), both lesion volume (β = −6.73) and tissue bridge width (β = −1.87) trended in the anticipated directions but did not reach statistical significance, most likely owing to the small sample size.

Scatterplots for all patients with follow-up data (**Figure 5, Panel A**) confirm that lesion volume and maximal axial damage ratio are significantly negatively associated with balance outcomes (r = −0.560, *p* = 0.030; r = −0.551, *p* = 0.033), whereas tissue bridge width showed a trend toward a positive association with dexterity (r = 0.454, *p* = 0.089). These relationships, however, were less robust in the post-surgical subgroup **(Figure 5, Panel B**), a finding that most likely reflects large interindividual variability and the lack of power in the study.

**Figure 5.**
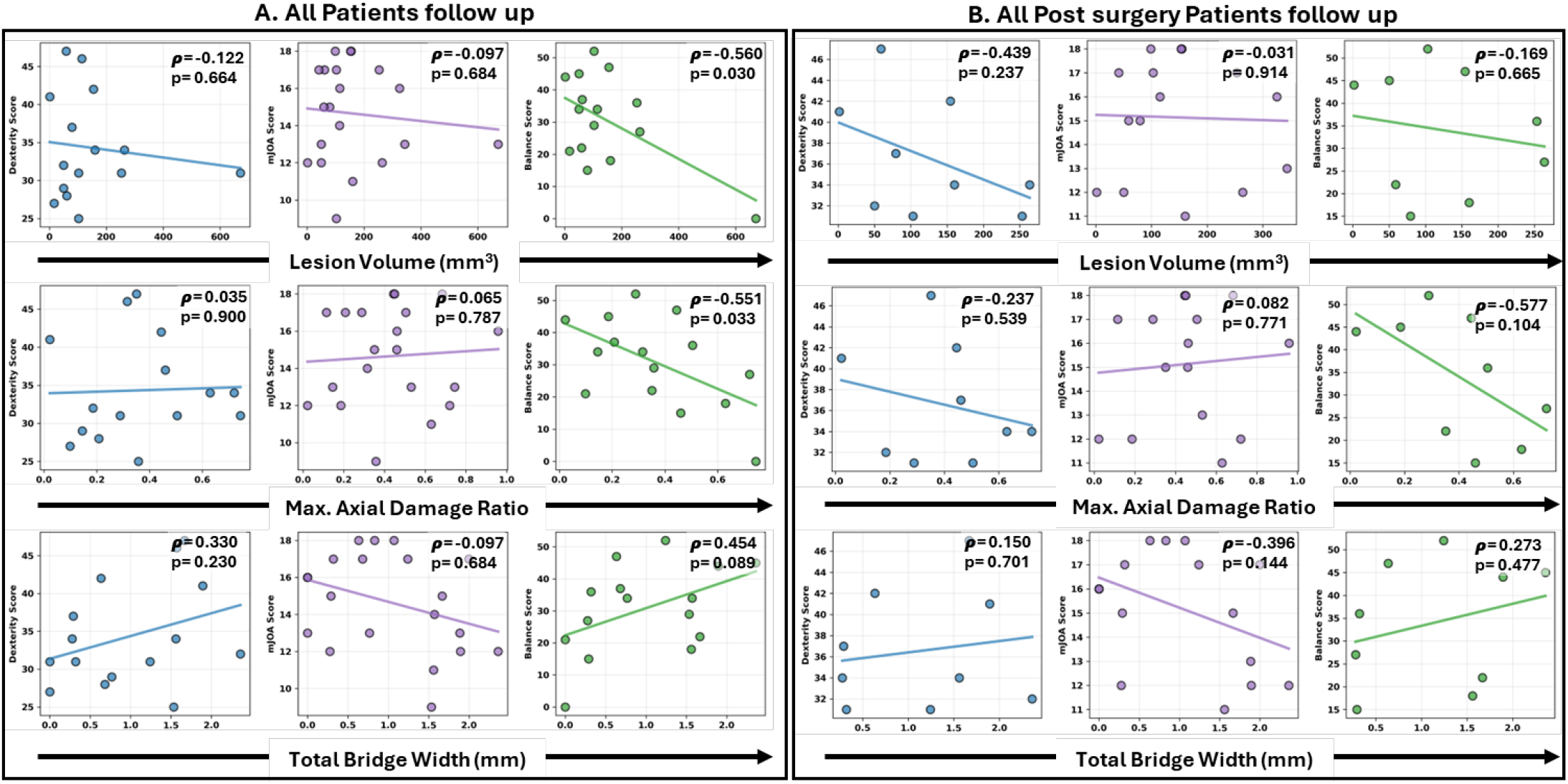
Partial correlation analysis between lesion characteristics and tissue bridges in patients with DCM with baseline and follow-up clinical outcomes. Partial correlation plots show the relationship between lesion volume, maximal axial damage ratio, and total bridge width with follow-up clinical scores, adjusting for baseline clinical scores. (A): All patients with a follow-up score regardless of surgery status (n =15–20): Top panel: Lesion volume was significantly associated with follow-up balance and trended toward an association with dexterity. Middle panel: Maximal axial damage ratio also showed a significant negative association with follow-up balance. Bottom panel: Total tissue bridge width showed trends toward positive associations with follow-up dexterity and balance but did not reach statistical significance. (B) Subgroup analysis in patients who underwent surgery (n = 9) revealed consistent trends: lesion volume and maximal axial damage ratio negatively associated with balance, and total bridge width positively associated with dexterity. Lines indicate the linear fit of the partial correlation models.

Overall, these findings suggest that lesion characteristics, particularly lesion volume and maximal axial damage ratio, are strong predictors of follow-up functional outcomes and that the preservation of tissue bridges may contribute meaningfully to upper-limb recovery in DCM.

## DISCUSSION

This study builds upon a growing body of research emphasizing the role of quantitative structural MRI features in delineating the pathology of DCM. Specifically, we focused on intramedullary T2w hyperintense lesions quantified by lesion volume, lesion length, maximal axial damage ratio, and injury-spared tissue bridges, which serve as measures of spinal cord damage that can be an imaging biomarkers of clinical severity and functional impairment^16,17^. Unlike prior studies relying on subjective manual segmentation, we employed SCIseg, a fully automated segmentation algorithm, applied across a multi-site DCM cohort^11,18,19^. This enabled reproducible and reliable lesion quantification across varying MRI scanners and clinical settings. Earlier work laid the groundwork of this study by manually characterizing lesion characteristics and surrounding tissue bridges, associating them with traumatic SCI outcomes^18^. We extended these findings to DCM as a non-traumatic SCI model and specifically evaluated their prognostic relevance in post-treatment recovery. Also, the use of automated segmentation can facilitate longitudinal tracking of lesion evolution offering a window into ongoing neurodegeneration or recovery following treatment. This dynamic monitoring capability can allow us to quantify changes in lesion burden of spared tissue over time, which could improve our understanding of DCM progression. Our results support the prognostic value of both intramedullary lesion burden and preserved tissue bridges, particularly in relation to hand dexterity and balance, hallmark dysfunctions in DCM. By utilizing automated, validated metrics, our study bridges the gap between manual assessments and scalable imaging biomarkers, laying the groundwork for personalized prognostic modeling in DCM.

In our cohort, higher lesion burden strongly correlated with DCM clinical severity. Patients with severe DCM showed significantly greater lesion volume, length, and maximal axial damage ratios compared to those with milder disease. These results extend existing literature linking lesion extent with neurological decline^18^. Further, our results are also consistent with the broader SCI literature, which implicates lesion burden in both structural compromise and secondary ischemic processes^20,21^. Notably, presence of lesion was associated with surgical rates, increasing the likelihood of decompression surgery even among those with mild disease, highlighting its role in clarifying treatment timing. These echoes work by Hilton et al., who found that MRI-based compression ratios explain surgical decisions in DCM^22,23^. Also, this aligns with acute SCI research showing lesion visualization to influence surgical management^24^.

Our study’s inclusion of multiple recruitment sites and surgeons ensured a diverse clinical setting and patient’s diversity, strengthening the generalizability of our findings. Quantitative lesion metrics including the number, volume, length, and maximal axial damage ratio were not only associated with baseline severity but also predicted long term deficits in dexterity and balance, key hallmark symptoms of DCM. These findings align with prior reports that T2w hyperintensity predicts poorer outcomes. Our study further extends this work by demonstrating that continuous lesion measures outperform binary lesion presence in prognostic models, collectively explaining 43% of the variance in follow-up clinical (balance) scores. By leveraging a fully automated, multi-site validated segmentation pipeline, our work provides a scalable framework to refine surgical timing and personalize prognostic estimates in DCM. Recent evidence shows that combined corticospinal and reticulospinal tract injury impairs mobility and increases spasticity, consistent with our finding that lesion metrics correlate with performance-based dexterity and balance but not with traditional subjective scores like mJOA or NDI^25^.

In a complementary analysis, we evaluated spinal cord tissue bridges – representing parenchymal neural tissue spanning the lesion – as a proxy for preserved axonal pathways. We found that wider tissue bridges were associated with better hand dexterity and balance scores, both at baseline and 6 months follow-up, whereas narrower or absent bridges correlated with poorer clinical scores. These findings support prior observations by Pfyffer et al., who reported that midsagittal tissue bridges after traumatic SCI predicted motor and sensory recovery and showed parallel increases with functional gains in both traumatic and ischemic injuries^7,8,26^. Further, Freund et al. hypothesized that such bridges may reflect spared intact axonal tracts and glial scaffolds supporting residual connectivity^7-9,18^. In our DCM cohort, patients with wider tissue bridges demonstrated significantly better recovery at 6 months. Given current limitations in tract-specific imaging under compression, region-based bridge analysis provides a feasible and informative surrogate for tract integrity. While the ventral corticospinal tract (CST) is often affected in DCM due to its midline location, tissue bridges on MRI may still represent spared or partially spared tracts located more dorsally or ventrally, rather than centrally^25^. According to Vallotton et al., preserved dorsal and ventral midsagittal neural tissues have been observed in both traumatic and degenerative injuries, even when the midline cord (where CST resides) is affected^27^. This suggests that tissue bridges may primarily represent dorsal column (sensory) and ventrolateral (reticulospinal and spinothalamic) pathways rather than central CST fibers. Similarly, Martin et al. (2022) emphasized that damage in DCM can be assessed through T1-weighted hypointense changes in MRI reflecting not only CST but also spinothalamic and dorsal column fiber pathology, indicating that non-CST tracts may contribute to preserved function when visible tissue bridges are present^28^.

Lesion burden and tissue bridge width were not mutually exclusive; they provided complementary insights with lesion size indicating structural compromise, while tissue bridge width suggested preserved function. Yet, uncertainty remains about the true integrity of axons within tissue bridges. In preclinical rat contusion models, sub-pial tissue bridges at the lesion center were found to be demyelinated between 4- and 7-days post-injury, rather than representing fully viable tracts^29^. Conversely, in clinical SCI cohorts, the width of tissue bridges at one-month post-injury predicted one-year motor recovery, supporting the notion that preserved tissue bridges may contribute to functional restoration^8^. Together, these data highlight that lesion architecture, particularly axonal disruption and demyelination, directly compromises the pathways critical for balance and dexterity. These observations are supported by recent work from Schading-Sassenhausen *et al*., who demonstrated that combined corticospinal and reticulospinal tract damage significantly impacts mobility and spastic muscle tone in SCI patients^25^. They underscore the need for integrative, multimodal assessments to also capture changes on molecular and microstructural levels.

From a pathological standpoint, intramedullary T2w hyperintense lesions on MRI reflect chronic destructive processes including gliosis, ischemia, and Wallerian degeneration that compromise both afferent and efferent pathways critical for motor coordination^30^. Larger lesions with greater axial involvement produce more extensive tract disruption, explaining their strong association with impaired dexterity and balance. In contrast, histological and preclinical studies demonstrate that these spared fibers enable functional reorganization and remyelination after injury. Mechanistically, our results suggest that tissue bridges may represent a surrogate for intrinsic repair potential and plasticity, potentially contributing to post-surgical recovery in DCM.

In our DCM cohort, lesion characteristics were strongly associated with objective measures of dexterity and balance yet showed no significant correlation with the mJOA or NDI scores. This disconnect suggests that quantitative imaging biomarkers can reveal structural pathology overlooked by symptom-based scales, and performance-based dexterity and balance tests may serve as more sensitive indicators of DCM pathology than traditional scales. Thus, mJOA and NDI, emphasizing more subjective clinician grading or patient-reported pain and motor symptoms, may underrepresent intramedullary damage.

While this study provided an automated assessment of intramedullary lesions and associated tissue bridges in a diverse DCM population, there are a number of limitations for consideration. First, our longitudinal cohort was modest in size, particularly in the post-surgical subgroup, which limited statistical power for treatment-specific analyses. Second, we evaluated only midsagittal tissue bridges; future studies should include full 3D spinal cord segmentation and parasagittal metrics for a more comprehensive assessment of spared tissue^31–33^. Third, electrophysiological measures were not included in this study, yet they could offer valuable complementary insight into functional integrity and tract-specific damage. Fourth, the follow-up period was limited to six months; however, recovery from DCM often extends over 12-24 months, and it is possible that the full effects of surgery were not captured at this interim time point. Longer-term studies are needed to assess the durability of tissue bridge morphology and its relationship to sustained clinical outcomes^34,35^. Fifth, although our automated pipeline performed robustly across sites and scanner platforms, variations in MRI field strengths and sequence parameters may still impact lesion and tissue bridge detection accuracy. Additionally, we did not explore why some DCM patients developed intramedullary lesions while others did not, a key question for understanding disease heterogeneity. In our cohort, greater lesion burden ^36^lected by larger volumes, greater lengths, and higher maximal axial damage ratios was strongly linked to more severe neurological impairment, though it remains unclear whether this reflects timing, onset, or progression of DCM. Finally, while tissue bridges may suggest structural preservation, they do not necessarily indicate neurophysiologically intact tracts. Preclinical models have shown early demyelination within tissue bridges, casting doubt on their functional viability. Yet, clinical studies consistently support tissue bridge width as a predictor of long-term recovery. These seemingly divergent findings emphasize that lesion architecture especially demyelination and tract disruption play a critical role in motor coordination loss and underscores the need for integrative, multimodal assessment.

In conclusion, by combining objective lesion metrics with sensitive measures of DCM clinical deficits, our approach offers a more precise framework for tailoring intervention timing and optimizing outcomes in DCM. Using a fully automated, multi-site segmentation pipeline, we derived high-resolution, reproducible measures of intramedullary lesion burden and tissue bridge architecture that reflect DCM pathophysiology and predict both clinical severity and postoperative recovery. Lesion volume, length, and maximal axial damage ratio emerged as negative predictors of hand dexterity and balance, while tissue bridge width served as a positive marker of preserved neural pathways and functional reserve. These complementary biomarkers could improve clinical decision-making, particularly for patients with mild or ambiguous symptoms, and support scalable, personalized prognostic modeling. In multivariable regression models, both lesion volume and maximal axial damage ratio predicted poorer outcomes, whereas tissue bridge width was a robust predictor of improved follow-up balance and dexterity. By demonstrating that continuous imaging metrics outperform binary classifications, our study advances the precision of DCM risk stratification. Clinically, incorporating these automated metrics across different MRI sequences and diverse scanner types could improve selection of patients at risk, inform surgical timing, and enhance outcome forecasting. Future studies should extend this framework to full 3D spinal cord segmentation, integrate electrophysiological and tract-specific imaging, and assess long-term functional durability. Together, these findings lay the foundation for precision-medicine strategies in DCM, enabling tailored therapeutic interventions and optimized recovery trajectories.

**Supplementary Figure 1.**
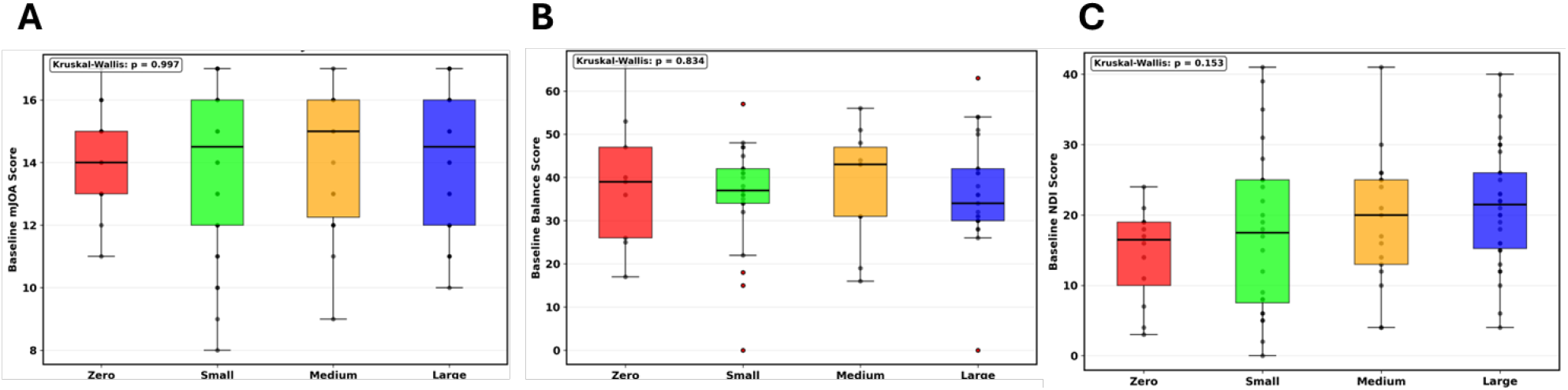
Group-wise comparison of baseline clinical scores and dexterity performance across varying degrees of bridge formation. Patients were categorized into four groups based on the extent of bridge formation: Zero - Bright Red, Small (<2.41 mm) - Bright Green, Medium (2.41mm to 5.18 mm) - Orange, Large (≥ 5.18 mm). (A) Baseline mJOA scores showed no significant differences across groups (Kruskal–Wallis, p = 0.997). (B) Baseline Balance scores also did not differ significantly (p = 0.834). (C) Baseline NDI scores trended toward significance but did not reach statistical threshold (p = 0.153).

**Supplementary Figure 2.**
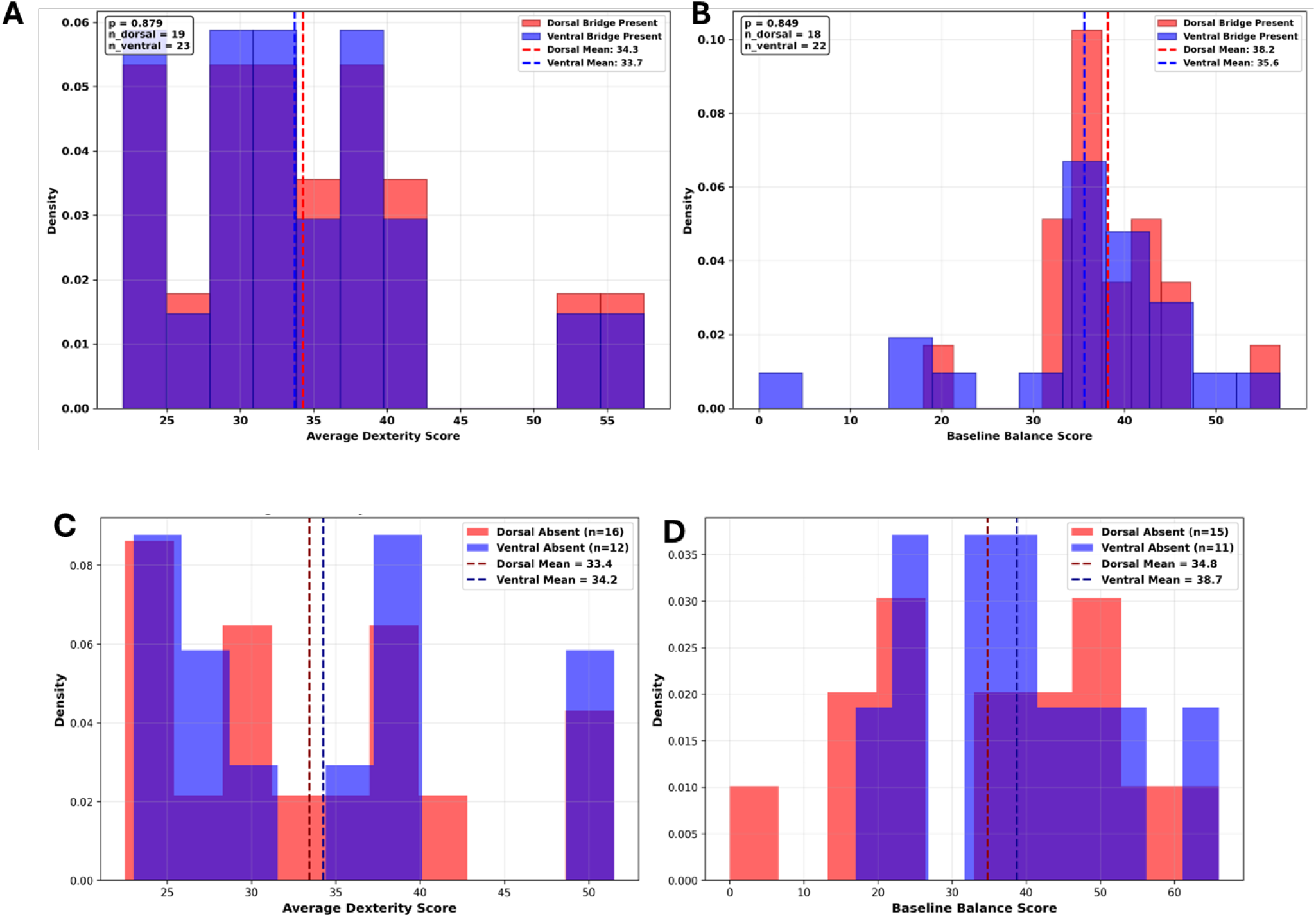
Distribution of clinical scores based on the presence or absence of dorsal and ventral spinal cord tissue bridges in DCM. Top Panels. Density plots comparing clinical scores in patients with preserved dorsal (red) versus ventral (blue) tissue bridges. A: Distribution of average dexterity scores. (B) Distribution of baseline balance scores. Vertical dashed lines indicate group means. Bottom panels: Density plots stratified by absence of dorsal (red) versus ventral (blue) bridges. C. Distribution of average dexterity scores for dorsal-absent (n = 16) and ventral-absent (n = 12) groups. D. Distribution of baseline balance scores for dorsal-absent (n = 15) and ventral-absent (n = 11) groups. Mean clinical scores are annotated with dashed lines. No statistically significant differences were observed across these conditions. These plots show that clinical scores are not significantly affected by the spatial presence or absence of specific tissue bridge types, although trends suggest slightly higher mean balance scores when dorsal bridges are preserved.

## Notes

**Funding:** This work was supported by Dr. Smith’s R01 grant from the National Institute of Neurological Disorders and Stroke (NIH/NINDS), award number R01 NS129852-01A1

### Competing Interest Statement

The authors have declared no competing interest.

### Funding Statement

This work was supported by R01 grant from the National Institute of Neurological Disorders and Stroke (NIH/NINDS), award number R01 NS129852-01A1

### Author Declarations

Standard Protocol Approvals, Registrations, and Patient Consents This retrospective, longitudinal, multi-site study included patients diagnosed with DCM across four tertiary academic centers: the University of Oklahoma Health Sciences Center (OUHSC), Northwestern University, University of Texas Southwestern (UTSW), and Stanford University. Institutional Review Board (IRB) approval was obtained from each participating institution, including OUHSC (IRB #15513). All patients provided written informed consent.

